# Metabolomic profiles of human glioma inform patient survival

**DOI:** 10.1101/2022.06.04.22275972

**Authors:** Andrew J. Scott, Luis O. Correa, Yilun Sun, Visweswaran Ravikumar, Anthony C. Andren, Li Zhang, Sudharsan Srinivasan, Neil Jairath, Kait Verbal, Karin Muraszko, Oren Sagher, Shannon A. Carty, Shawn Hervey-Jumper, Daniel Orringer, Michelle M. Kim, Larry Junck, Yoshie Umemura, Denise Leung, Sriram Venneti, Sandra Camelo-Piragua, Theodore S. Lawrence, Joseph E. Ippolito, Wajd N. Al-Holou, Prakash Chinnaiyan, Jason Heth, Arvind Rao, Costas A. Lyssiotis, Daniel R. Wahl

## Abstract

**Aims:** Targeting tumor metabolism may improve the outcomes for patients with glioblastoma (GBM). To further preclinical efforts targeting metabolism in GBM, we tested the hypothesis that brain tumors can be stratified into distinct metabolic groups with different patient outcomes. Therefore, to determine if tumor metabolites relate to patient survival, we profiled the metabolomes of human gliomas and correlated metabolic information with clinical data.

**Results:** We found that isocitrate dehydrogenase-wildtype (IDHwt) GBMs are metabolically distinguishable from IDH mutated (IDHmut) astrocytomas and oligodendrogliomas. Survival of patients with IDHmut gliomas was expectedly more favorable than those with IDHwt GBM, and metabolic signatures can stratify IDHwt GBMs subtypes with varying prognoses. Patients whose GBMs were enriched in amino acids had improved survival while those whose tumors were enriched for nucleobases and carbohydrates fared more poorly. These findings were recapitulated in validation cohorts using both metabolomic and transcriptomic data.

**Innovation:** Our results suggest the existence of metabolic subtypes of GBM with differing prognoses and further support the concept that metabolism may drive the aggressiveness of human gliomas.

**Conclusions:** Our data show that metabolic signatures of human gliomas can inform patient survival. These findings may be used clinically to tailor novel metabolically targeted agents for GBM patients with different metabolic phenotypes.

## Introduction

Glioblastoma (GBM) is the most common invasive primary brain tumor and nearly uniformly fatal despite surgical resection and standard of care chemoradiation. While therapeutic intervention has initial efficacy, tumors invariably recur and become resistant to treatment. Thus, there is an urgent need to identify and target molecular mediators of this resistance. While the therapy resistance and aggressiveness of GBM have been explored at genomic and transcriptomic levels, less is known about the metabolic mediators of therapy-resistant phenotypes.

Altered metabolism is a hallmark of cancers including GBM (1), and metabolic rewiring is critical for tumor cells to undergo conversion to aggressive and treatment-resistant phenotypes. Tumor metabolism is influenced by both cancer cell-intrinsic information (genome, epigenome, proteome, post-translational modifications) and cell-extrinsic cues from the tumor microenvironment. Given that targeting metabolism has a history of success in a variety of cancers, targeting the metabolic phenotypes of GBM cells may represent an effective treatment strategy (2). Indeed, early data from several metabolically targeted therapies for GBM patients have yielded promising outcomes (3, 4).

Understanding the metabolic phenotypes of gliomas could also provide information about tumor aggressiveness and patient prognosis. Altered expression of metabolic enzymes or imaging-defined glucose uptake can inform prognosis in a variety of cancers including glioma (5-8). Metabolite levels themselves can distinguish low grade and high-grade gliomas and suggest that GBMs favor anabolic metabolism and heterotrophy (9, 10). Whether these metabolomic profiles can provide information regarding glioma patient outcome remains uncertain.

To address this question, we measured the metabolomes of 69 patient gliomas and found that tumors robustly cluster into categories representing isocitrate dehydrogenase-wildtype (IDHwt) GBM and IDH-mutant (IDHmut) gliomas based on metabolic profiles. IDHmut gliomas further separate high grade (grade 4 astrocytoma) from lower grade tumors (grades 2 and 3 astrocytomas and oligodendrogliomas). Further analyses of GBM tumors reveals distinct metabolic subtypes with different patient survival times. We found no relation of these subtypes to known survival predictors, suggesting metabolism can influence GBM progression independently of these factors. Taken together, these findings suggest the existence of discrete metabolic GBM subtypes and may pave the way for therapies targeting metabolic pathway activity to improve patient outcome.

## Results

### Metabolomic profiling distinguishes IDH-mutant from IDH-wildtype gliomas

Using the University of Michigan Brain Tumor Bank, we identified 69 flash-frozen glioma samples with sufficient tissue for metabolomic analysis. All samples were deemed to contain 70% or greater viable tumor content at time of resection after quality assurance by a clinical neuropathologist (S.V. and S.C-P.). Clinical data associated with these tumor samples was then obtained from the medical record. This cohort (**Table 1, Figure 1**) contained IDHmut oligodendrogliomas (29%), IDHmut astrocytomas (12%), and IDHwt GBMs (59%), all of which were molecularly defined using the 2021 WHO criteria (11). Overall median survival, sex ratios, and MGMT promoter methylation status of all three groups were as expected with median survival times of around 11 years for IDHmut oligodendrogliomas, 7 years for IDHmut astrocytomas and 1.6 years for IDHwt GBMs (12). All patients were treated with some extent of resection (as opposed to biopsy-alone) due to the requirement of sufficient tissue for banking. Within this cohort, 63% of patients with IDHmut astrocytomas, 67% of patients with IDHmut oligodendrogliomas, and 95% of those with IDHwt GBMs received both RT and chemotherapy, typically temozolomide, at some point following resection.

**Table 1.** Patient Characteristics. Resected tumor samples stored in the University of Michigan Brain Tumor Bank were matched with patient medical records to determine the indicated information. Age and performance status are shown as median ± interquartile range.

|  |  |  |
| --- | --- | --- |
| Total cohort n=69 |  |  |
| IDH mutant oligodendroglioma, n=20 |  |  |
|  | Age | 38.6 ± 17.5 |
|  | Sex | Male 55%, female 45% |
|  | Extent of resection | GTR 50%, NTR 50% |
|  | Performance status | 1.0 ± 0.75 |
|  | Receipt of radiation | 75.0% |
|  | Receipt of alkylating chemotherapy | 77.8% (14/18, 2 unknown) |
| IDH mutant astrocytoma, n=8 |  |  |
|  | Age | 34.0 ± 15.0 |
|  | Sex | Male 50%, female 50% |
|  | Extent of resection | GTR 25%, NTR 75% |
|  | Performance status | 1.5 ± 1.8 |
|  | MGMT methylation status | Methylated 57.1% |
|  | Receipt of radiation | 62.5% |
|  | Receipt of alkylating chemotherapy | 62.5% |
| IDHwt GBM, n=41 |  |  |
|  | Age | 59.7 ± 13.3 |
|  | Sex | Male 62.5%, female 37.5% |
|  | Extent of resection | GTR 53.7%, NTR 34.1%, STR 12.2% |
|  | Performance status | 1.0 ± 1.0 |
|  | MGMT methylation status | Methylated 36.6% |
|  | Receipt of radiation | 97.6% |
|  | Receipt of alkylating chemotherapy | 95.1% |

**Figure 1.**
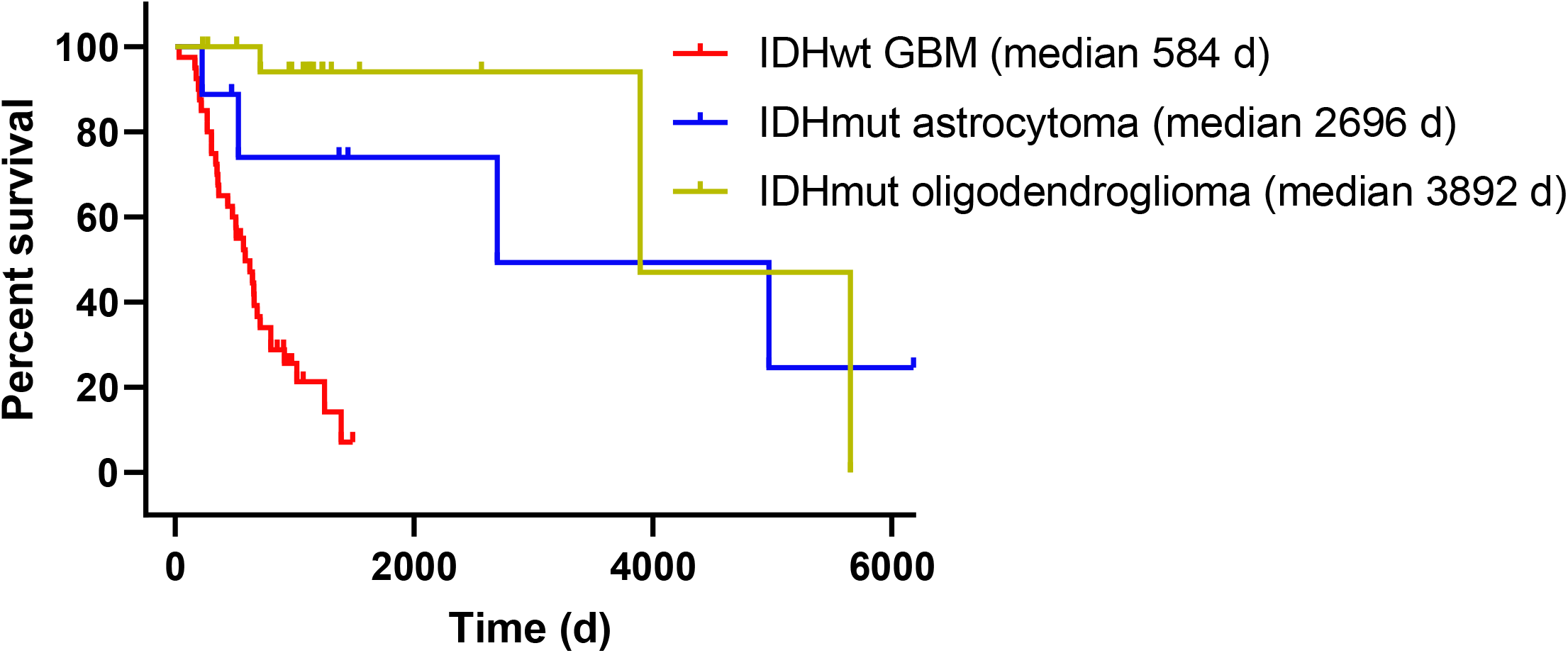
Overall survival times for glioma patients. Kaplan-Meier curves of overall survival times for glioma patients corresponding to tissue samples acquired from the University of Michigan Brain Tumor Bank with the indicated tumor types are shown. Patients with unknown survival times were censored at time of last follow-up.

We first asked whether metabolomic information from our tumor samples would be of sufficient quality to discriminate between known biologic tumor types. Within this cohort and for each tumor sample, we extracted polar metabolites and performed quantification by liquid chromatography-mass spectrometry (LC-MS) as described previously (13). With this method we determined relative abundances of over 200 compounds comprising central carbon, nucleotide, and amino acid metabolism. To visualize whether our high dimensional metabolite information was sufficient to group tumor samples based on original identity, we performed uniform manifold approximation and projection (UMAP). UMAP analysis revealed that IDHwt GBMs tend to separate from IDHmut gliomas, while IDHmut subsets (astrocytoma and oligodendroglioma) are more metabolically similar (**Figure 2A**).

**Figure 2.**
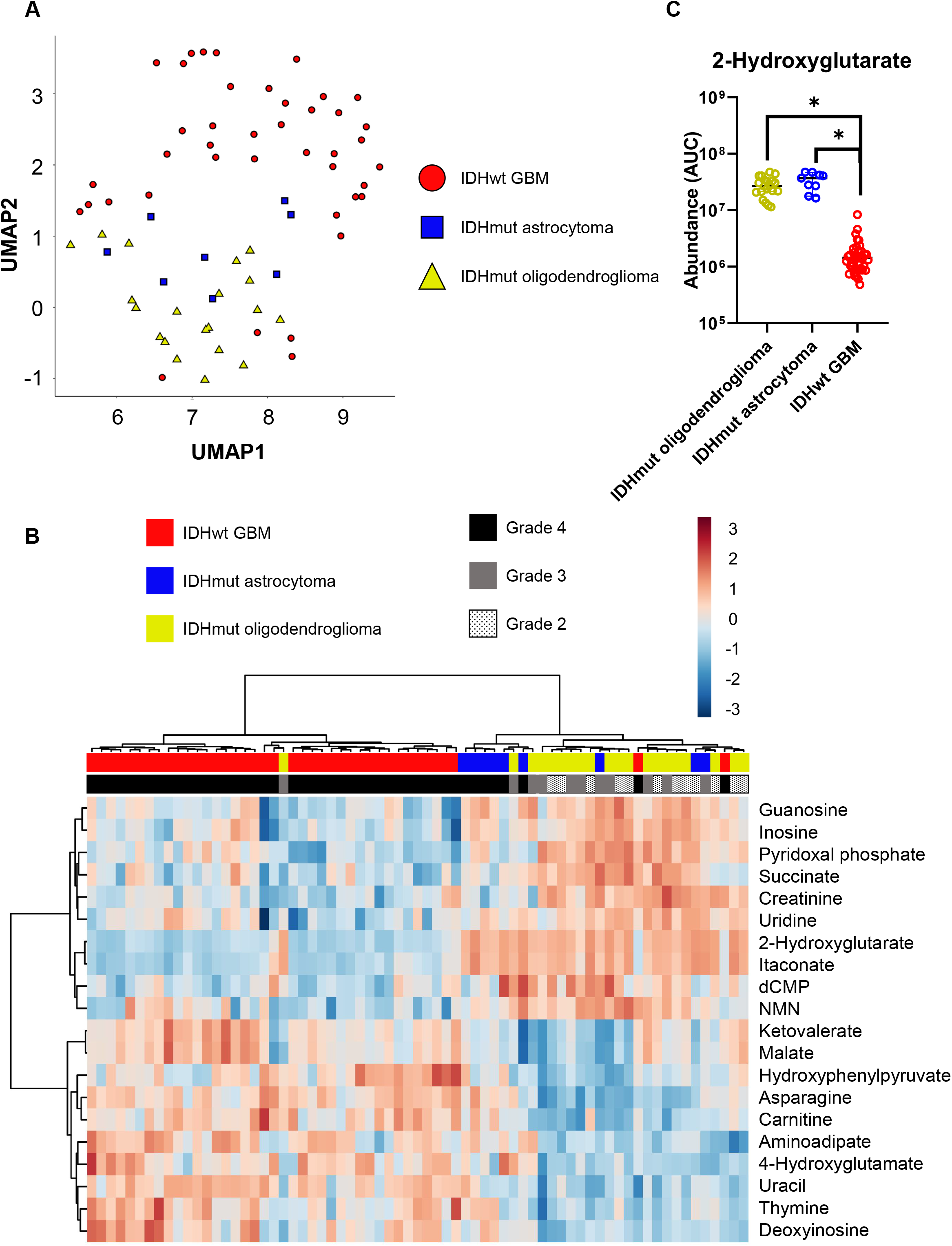
Metabolomics distinguishes IDHmut from IDHwt gliomas. *A*, Metabolite levels of tumors from glioma patients were measured by LC-MS and then assessed by UMAP. *B*, Metabolite levels were assessed by data reduction with Binner followed by unsupervised hierarchical clustering. Color scale indicates log-transformed values of data points after normalization to the median area under the curve (AUC) of each compound. *C*, Levels of 2HG were determined by measuring LC-MS AUCs matched to ion fragmentation data.

When levels of tumor metabolites across patients were analyzed by unsupervised hierarchical clustering, two distinct metabolomic groups were immediately apparent, with one cluster representing IDHwt GBM and the other representing IDHmut astrocytoma and oligodendroglioma (**Figure 2B, Supplemental Figure 1**). This is consistent with our UMAP analysis (**Figure 2A**). Mutations of IDH, typically at an arginine residue required for substrate recognition, cause an accumulation of 2-hydroxyglutarate (2HG, (14-17)). As expected, levels of 2HG were 10 to 50-fold higher in IDHmut tumors than IDHwt GBMs (**Figure 2C**). Surprisingly, our method identified several other metabolites, including itaconate, citramalate and ketoleucine, that were elevated to a similar magnitude as 2HG in IDHmut tumors (**Supplemental Figure 1**). These findings could hint at interesting new biology in IDHmut gliomas, or they could be mis-called metabolites due to their chemical similarity to 2HG (*e*.*g*., similar fragmentation patterns and retention times on chromatography). To discriminate between these possibilities, we processed our data using Binner (18), which identified isobaric overlaps of 2HG with citramalate and itaconate with ketoleucine. Following this additional processing of LC-MS data, IDHwt GBMs remained distinct from IDHmut gliomas (**Figure 2B**). Together, these data show that our tumor metabolomic data is of sufficient quality to discriminate known biologic subtypes of glioma and would allow for deeper investigations for novel biology.

Grade 4 IDHmut astrocytomas have a worse prognosis than grade 2 or 3 IDHmut astrocytomas, but the prognostic difference between grade 2 and 3 IDHmut gliomas is uncertain in the era of molecularly defined tumors (19). Notably, grade 4 IDHmut astrocytomas clustered together, and lower grade 2 and 3 tumors remained intermixed on the basis of metabolites (**Figure 2B**). While IDHmut grade 4 astrocytomas had similar levels of 2HG as lower grade IDH mutant tumors, their levels of asparagine and several other metabolites were more similar to IDHwt GBMs than to lower grade IDHmut tumors. Hierarchical clustering could not discriminate between grade 2 or grade 3 IDHmut tumors (**Figure 2B**).

We also noted that two IDHwt GBMs clustered with low-grade IDHmut tumors (**Figure 2B**). This clustering was not due to alternative IDH mutations missed by immunohistochemistry, as 2HG levels were similar to other IDHwt GBMs. Rather, these two tumors had similar levels of succinate, creatinine and other metabolites as the IDHmut tumors. Additional investigation of these two unusual GBM cases found survival times substantially longer than the 1.5-year median for GBM, similar to IDHmut gliomas. One patient in their early fifties survived 4 years beyond diagnosis and the other (early twenties, far below median age of 65 years) is still alive 5.5 years after diagnosis at the time of writing. A single IDHmut grade 3 oligodendroglioma metabolically clustered with IDHwt GBMs. This tumor was one of only 3 recurrent oligodendrogliomas in our cohort, and this patient survived 2.5 years following re-resection, suggesting more aggressive behavior than a typical oligodendroglioma.

### Metabolomics-based clustering bins GBM patients into groups with different prognoses

The data above confirmed that IDHwt GBMs have a metabolic phenotype distinct from IDHmut gliomas and suggested that our data was of sufficient quality to investigate less understood metabolic pathways in glioma. Outcome for GBM is dramatically worse than for IDH mutant tumors. This poor survival rate may be at least partially due to metabolic phenotype (2, 9, 20), and we and others have demonstrated that targeting metabolism in GBM can improve survival in animal models and is under investigation in patients (4, 21, 22). To determine if tumor metabolomic profiles are related to patient survival in GBM, we questioned if GBMs can be grouped into different metabolic subtypes with different survival times in a manner similar to efforts to categorize GBMs by transcriptomic and DNA methylation patterns (23, 24).

We first confirmed our GBM tumor samples reflected a typical clinical cohort by univariate analyses with known survival factors. As expected, older age, male sex, poor performance status, and an unmethylated MGMT promoter were all associated with inferior survival within this cohort of GBM patients (**Supplemental Table 1**), though some variables did not achieve statistical significance. We then performed unsupervised hierarchical clustering of metabolites in the 41 tumors from GBM patients. This analysis identified 3 unique metabolic clusters of GBMs (**Figure 3A**). Inspection of individual metabolites represented by each subtype revealed enrichment of either 1) nucleobases and carbohydrates (Base/Carb), 2) amino acids (AA), or 3) metabolites associated with oxidative stress regulation and mature nucleoside/nucleotide species (Nuc/Ox).

**Figure 3.**
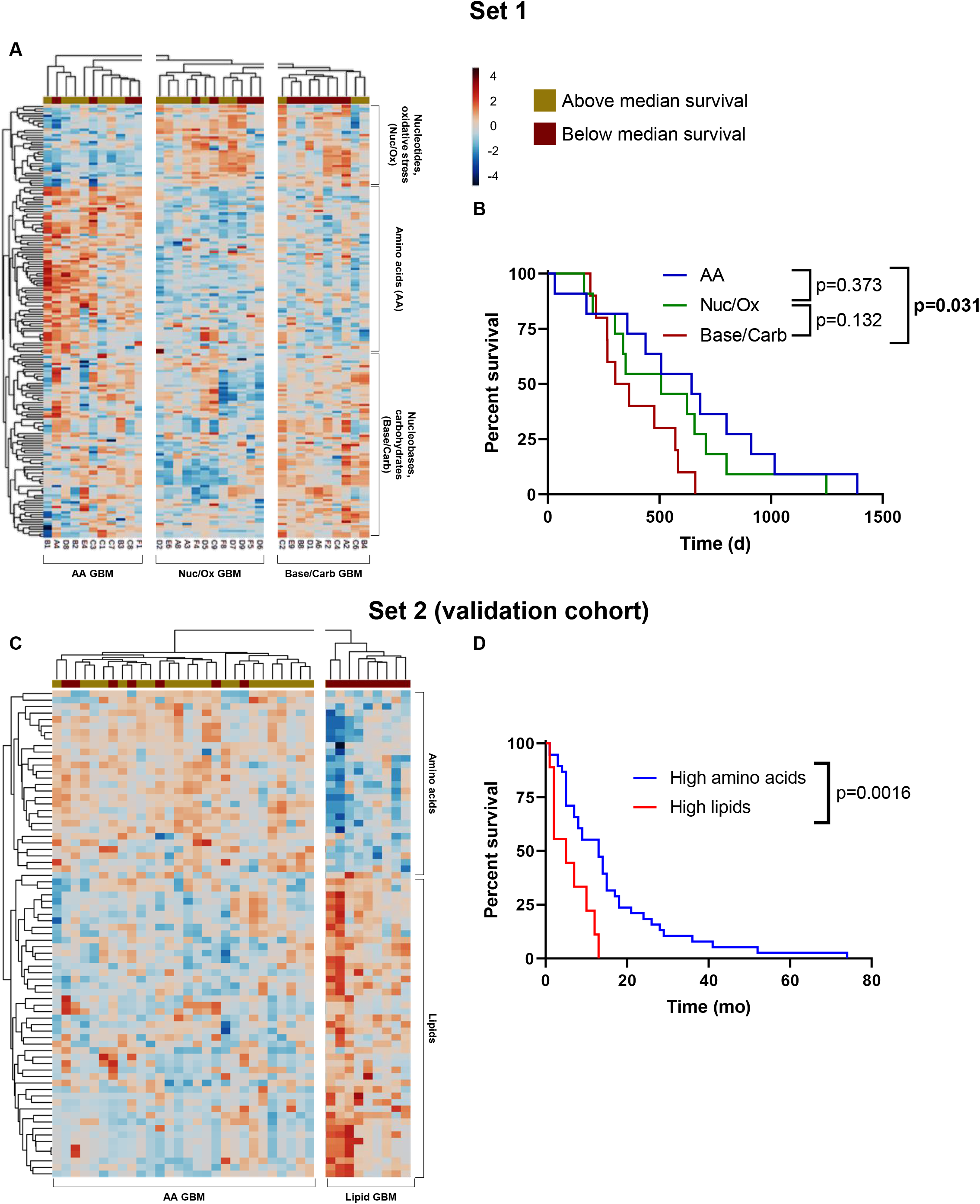
Metabolomics-based clustering bins GBM patients into groups of different prognosis. *A*, Levels of metabolites in GBM patients with known survival times were organized by unsupervised hierarchical clustering. *B*, Kaplan-Meier curves with survival times for patients in the subtypes identified in panel *A. C*, To validate findings in panels *A* and *B*, a second, independent cohort was assessed using different mass spectrometry methods and then analyzed by clustering as in panel *A. D*, Kaplan-Meier curves with survival times for patients in the subtypes identified in panel *C*. Color scales for both heatmaps indicate log-transformed values of data points after normalization to the median AUC of each compound.

Among these three putative metabolic subgroups of GBM, the Base/Carb group was predominantly comprised of patients with below-median survival, and GBM patients with above-median survival tended to fall within the AA and Nuc/Ox clusters (**Figure 3A**). Survival analysis of these 3 groups confirmed prognostic differences, with Base/Carb patients showing the worst survival and the AA group showing superior survival (**Figure 3B**). These differences were not due to receipt of different treatments, as receipt of RT and temozolomide was not different between groups (**Supplementary Figure 2A,B**). These findings are reminiscent of our previous work showing that GBMs with high levels of nucleotides and their derivatives, particularly purines, are especially resistant to treatment (21).

We then asked if survival differences among metabolic subtypes were related to known clinical predictors of GBM patient survival. Assessment of established predictors of GBM patient survival (sex, age at diagnosis, MGMT promoter methylation, performance status and extent of resection) within each subtype determined a mostly even distribution of these factors across metabolic groups (**Supplemental Figure 2C-G**). MGMT promoter methylation, despite being a predictor of longer survival, was notably higher in the poor-prognosis Base/Carb group than the AA and Nuc/Ox groups. Taken together these observations indicate that the metabolic features of GBM patient tumors might provide information regarding patient prognosis that could complement the information conveyed by conventionally used clinical information.

To validate our findings that GBMs can cluster into metabolic groups with different prognosis, we assessed a second, independent dataset (validation cohort) of GBM tumor specimens containing both metabolomic profiles and survival times (9, 20). While metabolomic data from the validation cohort contained largely different metabolites due to different LC-MS detection methods, patients in the validation cohort could also be binned into metabolically defined subtypes with differing prognosis that were similar to those found in our initial cohort of tumors (**Figure 3C**). Consistent with the data obtained using our method, an amino acid-high subtype with superior survival was identified (**Figure 3D**). The second subtype within our validation cohort contained high levels of many lipid species and had a significantly reduced median survival compared to the amino acid-high subtype. The identification of a lipid-high subtype in this dataset, rather than the Base/Carb and Nuc/Ox groups reported in set 1, is likely due to differences in LC-MS compound detection methods with limited detection of nucleobases that are increased in patients with worse survival.

### Association of individual metabolites with GBM patient survival

Having identified metabolic signatures that correlated with GBM patient survival, we next explored if patient outcome was related to levels of individual metabolites. For each metabolite in each independent dataset, we determined hazard ratios (**Supplemental Figure 3A-B**) and correlation with survival (**Supplemental Figure 3C-D**). While a variety of metabolites exceeded the 95% confidence interval, no singular metabolite appeared to reliably predict survival with a p-value of <0.05 in both sets. This may be due to sample quality, different analysis methods across data sets, or the inherently dynamic nature of metabolite levels.

We then asked if patients with below-median vs. above-median survival were metabolically distinguishable when the two groups were directly compared. To this end, we used partial least squares discriminant analysis (PLS-DA) to identify metabolic features that could discriminate between above median survivors and below median survivors (**Figure 4A,B)**. In this comparison we found that levels of purines including AMP/dGMP and adenine were significantly elevated in patients with inferior survival, as well as a variety of other purine and pyrimidine metabolites (**Figure 4C**). This agrees both with our metabolic clustering (**Figure 3**) and with our previous data showing that purines promote therapeutic resistance in GBM (21). When we assessed our validation cohort, we found several diverse lipid and amino acid species were different between groups, in agreement with our metabolic clustering analysis (**Figure 4D)**. Distinct from these two metabolite categories, levels of ascorbate were notably higher in above-median than below-median survivors, in agreement with ongoing clinical strategies to modulate ascorbate levels in GBM patients to improve outcome (3, 22).

**Figure 4.**
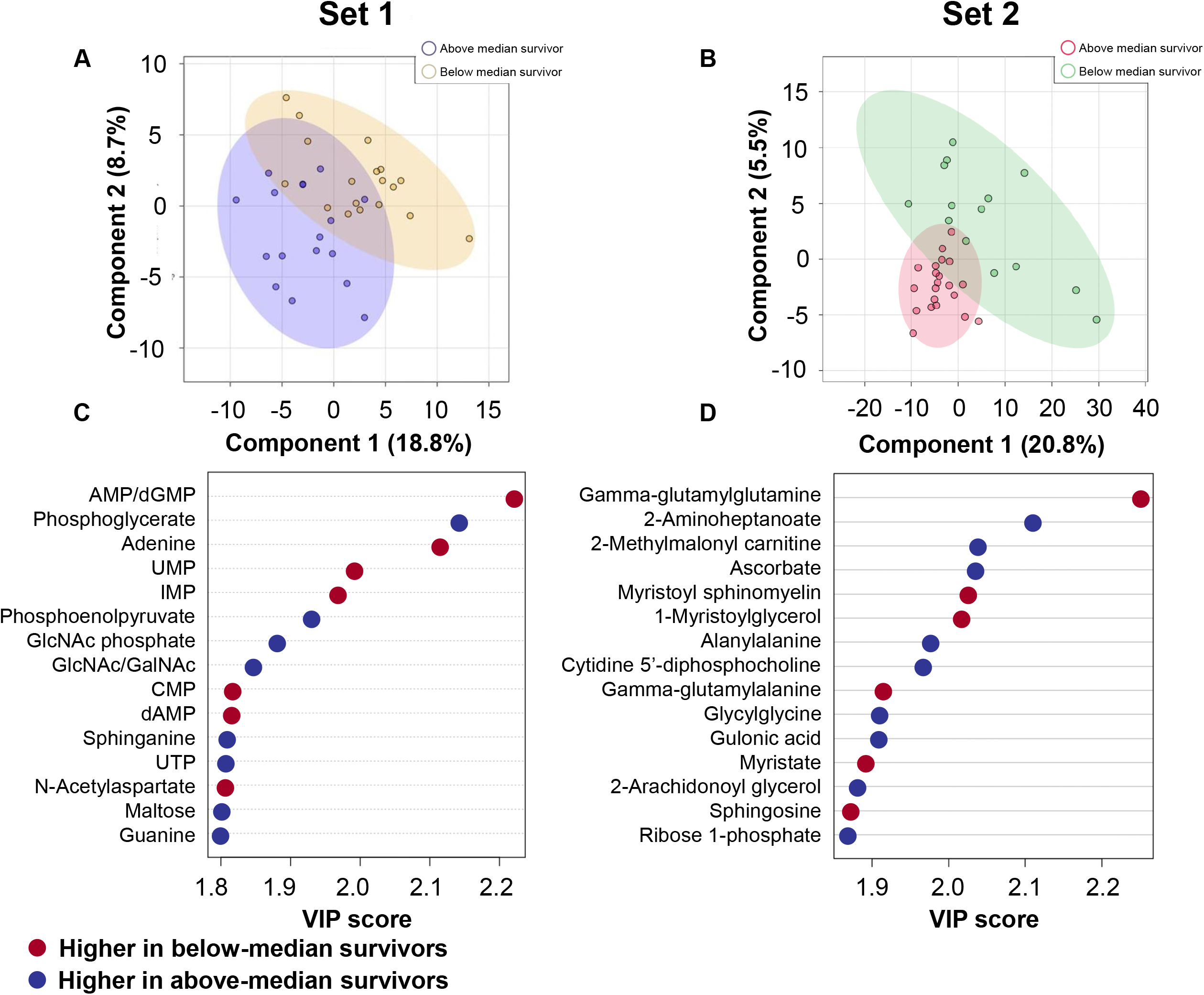
Assessment of individual metabolites in patients with different outcomes. *A-B*, PLS-DA of metabolomics datasets. *C-D*, VIP scores for metabolites were determined in tumors from below-median vs. above-median survivors.

### Metabolomic analysis of primary vs. recurrent GBM

Surgical resection and standard chemoradiation therapy improve survival for GBM patients, but this initial efficacy is limited by the development of treatment resistance. Recurrent, therapy-resistant tumors develop within the high-dose radiation field, and the ability of recurrent tumors to resist therapy is in part due to metabolic alterations within the tumor (2). Therefore, we asked if recurrent GBMs were metabolically distinct from primary GBM. Metabolomic profiles between primary and recurrent GBM tumor samples were largely similar with respect to central carbon and amino acid metabolites. In contrast, we observed differences in individual metabolites that agree with their known roles in tumorigenesis. Levels of guanosine, which may promote glioma stemness, gliomagenesis and treatment resistance (7, 21, 25, 26), were approximately 50% higher in recurrent tumors than in primary GBM (**Figure 5A**). Decreased levels of cystathionine (a precursor to glutathione) and aconitate (a TCA cycle intermediate) were also observed in recurrent GBM compared to primary GBM (**Figure 5B-C**). To further determine if recurrent GBM is metabolically distinguishable from primary GBM, we performed metabolite set enrichment analysis (MSEA) of compounds whose average abundance differed by more than 50% between groups. Metabolite sets most enriched in recurrent GBM agreed with our survival analysis. That is, we observed pathways that we and others have linked to patient outcome and tumor progression, including carbohydrate, nucleotide, tryptophan and ascorbate metabolism (9, 20-22), which were further enriched in recurrent GBM (**Figure 5D**) and suggest these may be promising leads to target metabolic activity in patients with recurrent GBM.

**Figure 5.**
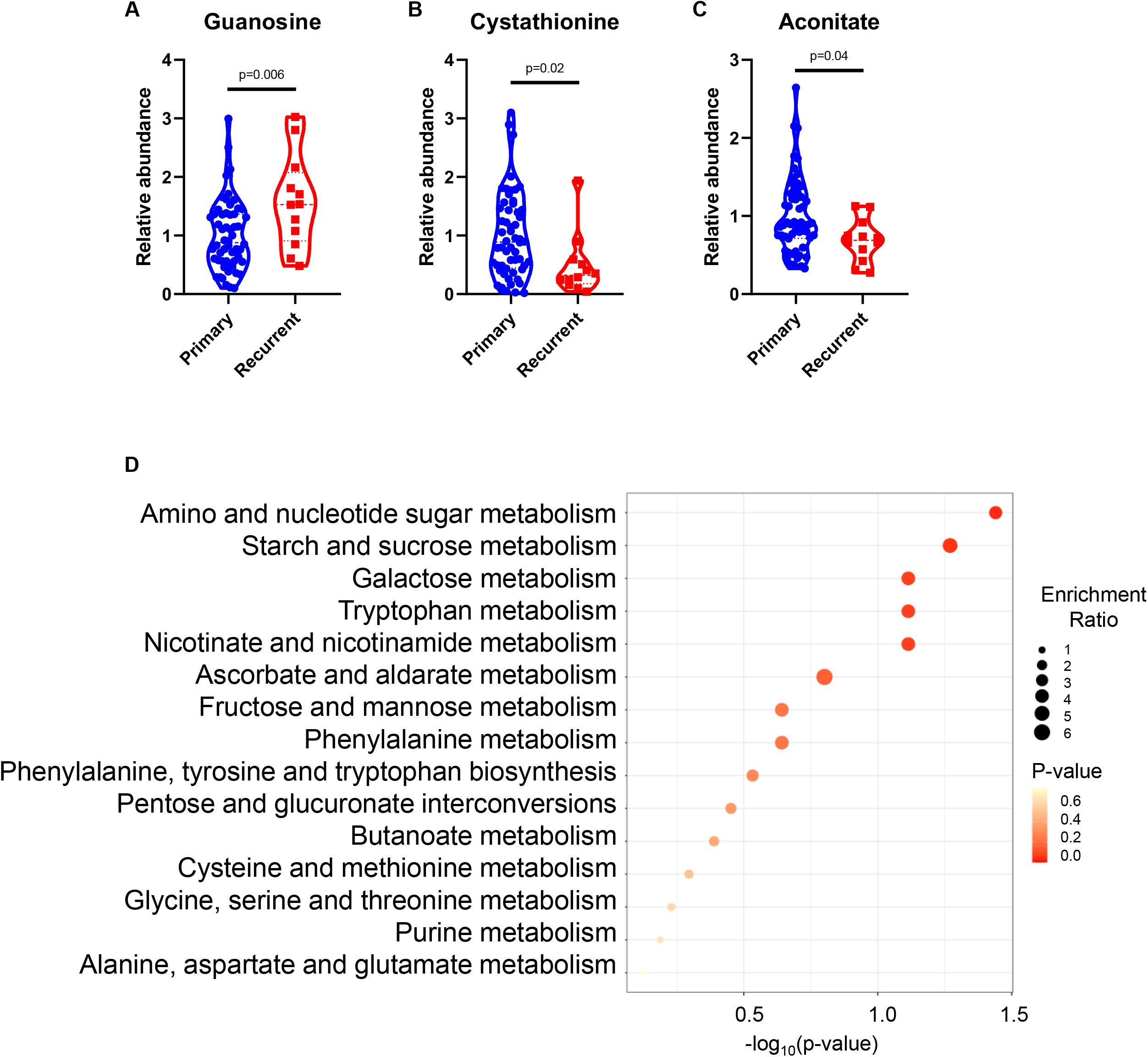
Metabolomic analyses of primary vs. recurrent GBM. *A-C*, Relative metabolite abundances in primary vs. recurrent GBM. *D*, MSEA of metabolites differing by more than 50% between primary and recurrent GBM.

### Validation of metabolic subtypes by transcriptomic analysis

Metabolomic analysis of brain tumors is not a standard part of clinical care, in part due to the logistical challenges of quickly flash-freezing tumor tissue and the cost of metabolomic analysis. We wanted to understand if our metabolism-centric approach to understanding GBM patient outcomes could translate into standard clinical settings where approaches such as exome sequencing and transcriptomic analysis are more commonly performed. Metabolites are linked by the enzymes that catalyze their interconversion, and the levels of these enzymes are quantified in transcriptomic analyses such as RNAseq.

We questioned if it were possible to validate metabolic GBM subtypes and their prognostic utility at the transcriptional level. Therefore, we selected the metabolites of the compound-high clusters characterizing each subtype and performed joint pathway analysis with MetaboAnalyst 5.0 (27) for each set. This approach allowed us to identify connections globally across gene-metabolite networks and predict the genes likely involved in the metabolic activities of each subtype (**Figure 6A-C**). We first defined a gene set for each tumor type (Base/Carb, Nuc/Ox and AA) comprised of the genes with at least two connections to respective input metabolites. Thus, these sets of genes were predicted to contribute to tumor metabolic phenotype. We then interrogated linked expression and outcome data from the TCGA to determine how these gene sets were associated with GBM patient survival. For each transcriptional signature, TCGA GBM tumor samples were scored using ssGSEA (28) and then assessed for survival in high-scoring (>0) vs. low-scoring (<0) groups. Similarly, we performed separate analyses of patient survival in each metabolite-defined subtype vs. the remainder of the cohort.

**Figure 6.**
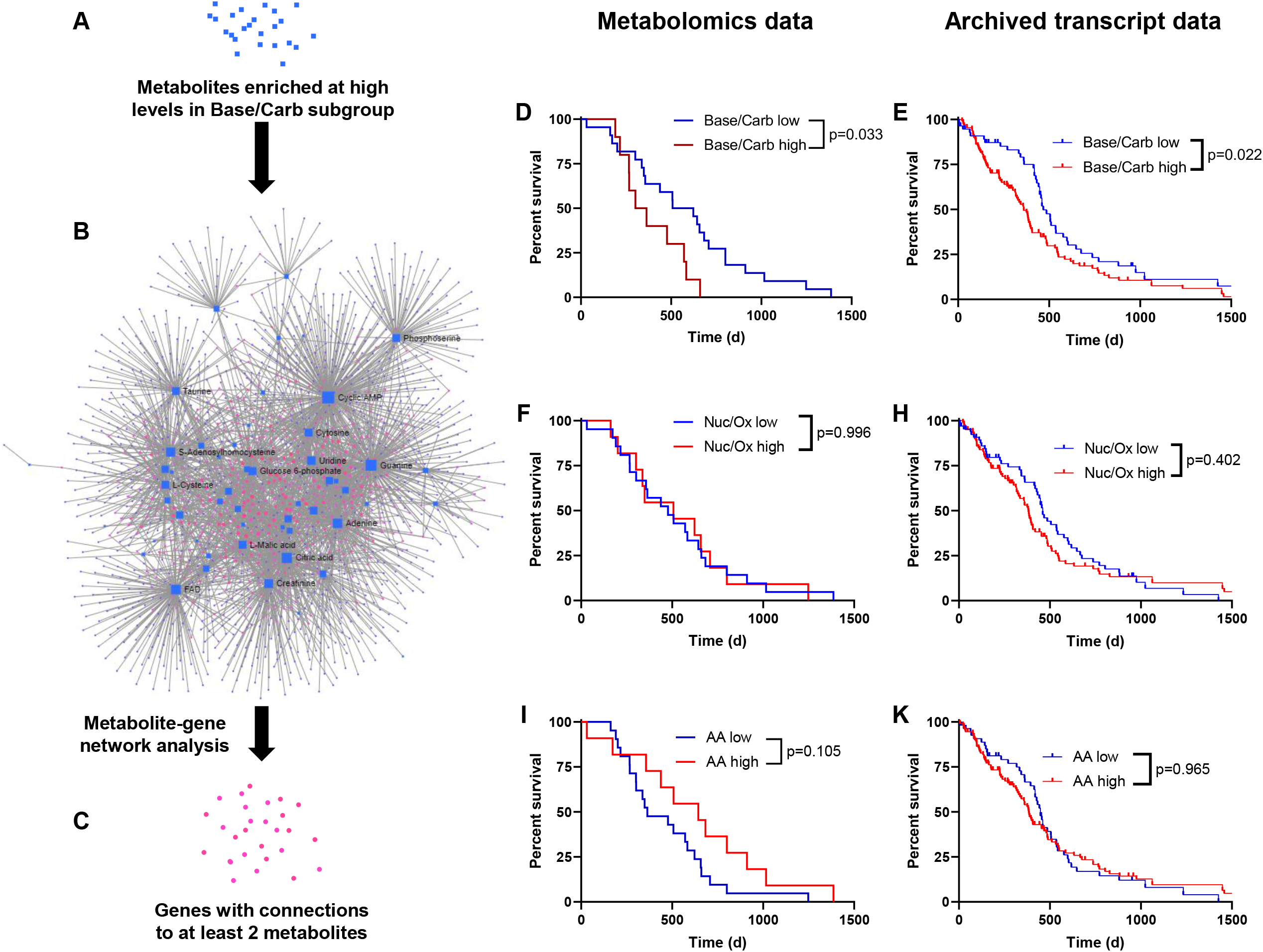
Transcript validation of GBM metabolic subtypes. *A*, Metabolites representing each metabolic GBM subtype were used to identify relevant RNA transcripts by metabolite-gene interactions with high confidence. *B*, Network of genes and metabolites from the Base/Carb GBM subtype. Blue squares represent metabolites; magenta circles represent genes. *C*, Genes from panel *B* with connections to at least 2 metabolites were selected for transcriptomic analysis. *D, F*, I, Kaplan-Meier curves with patients from the indicated metabolic subtypes vs. the remainder of the cohort. *E, H, K*, Kaplan-Meier curves using archived transcriptomic data with patients from high vs. low transcriptomic scores corresponding to the genes identified for each subtype.

Comparison of survival curves between metabolite-defined groups and corresponding transcriptionally defined groups showed close agreement. Patients in the Base/Carb subtype had inferior survival (**Figure 6D**), as did those in the transcriptionally defined Base/Carb group (**Figure 6E**). Analysis of both the metabolically and transcriptionally defined Nuc/Ox groups were also consistent, with neither showing differences in survival from the rest of the cohort (**Figure 6F-H**). While the metabolically defined AA group trended with a higher median survival, the transcriptional AA group showed no significant difference in survival from the rest of the cohort (**Figure 6I-K**). Collectively, these data suggest that the Base/Carb subtype can be transcriptionally distinguished from the other metabolic subtypes of GBM.

## Discussion

In this study we have nominated metabolomic subtypes for GBM that inform patient prognosis and could lead to new treatment strategies. IDHwt GBMs are metabolically separable from IDHmut gliomas, and within the IDHmut group of gliomas grade 4 astrocytomas are metabolically distinct from grade 2 and 3 gliomas. IDHwt GBM and IDHmut gliomas also differ dramatically by 2HG levels, with IDHmut gliomas containing expectedly higher levels than IDHwt GBMs by up to 50-fold. GBMs can be further separated into metabolic groups with different survival times that agree with transcriptional analysis of metabolite-associated genes. These data indicate that metabolic phenotypes of brain tumors may be able to inform patient outcome and tumor aggressiveness.

There are several general reasons metabolite levels could contribute to patient outcome. Metabolites are likely to regulate therapeutic responses to radiation and temozolomide, which are the predominant treatments prescribed for GBM. For example, high nucleobase levels could provide a readily accessible pool of substrates for nucleic acid synthesis required for proliferation, and/or the production of mature nucleosides and nucleotides that mediate radiotherapy resistance (21). High levels of carbohydrates might similarly feed anabolic pathways contributing to tumor aggressiveness (9). We also observed that tumors rich in lipids were linked to worse patient outcome. This can be explained by numerous cellular activities for lipids, including membrane production in proliferating cells, second messenger activity for proliferative and survival signaling pathways, and oxidation in the mitochondria to produce redox cofactors and ATP (29, 30).

In contrast to carbohydrates, nucleotide species, and lipids, levels of amino acids were associated with more favorable outcomes. Notably, synthesis and degradation pathways of amino acids are more diverse, branching about numerous metabolic pathways. Thus, GBMs with high amino acid levels might represent a more globally active network of metabolic pathways without reserve capacity and thus may be more easily perturbed by therapy. Alternatively, or in addition, high amino acid levels might also reflect lower levels of protein synthesis. Both scenarios could in turn could lead to more favorable survival. Our observation that predicted genes representing the AA subtype did not correlate with survival (in contrast to metabolite levels) could be explained by several hypotheses. Amino acid pathways represented by the indicated metabolites and transcripts might be regulated post-transcriptionally, in which case transcriptional data would not represent actual metabolic activity. Alternatively, or in addition, metabolite levels do not necessarily reflect pathway activity; for example, high amino acid levels could arise either from increased production or decreased utilization.

At present, metabolomic analysis of patient tumors is infrequently performed in clinical practice. Flash freezing tumor specimens immediately after resection and subsequent metabolomic analysis is logistically difficult and costly. There is also a lack of standardization regarding metabolomic detection methods and analysis across academic and medical centers. We directly observed this difficulty in the different types of metabolites detected between our initial and validation cohorts, which were generated using different platforms. However, further investigation of the metabolic underpinnings of GBM or other cancers might lead to standardized diagnostic and prognostic methods for metabolite detection and quantification. Studies employing analysis of transcriptional or proteomic signatures with patient-matched metabolomic data could identify molecular profiles that directly correspond to metabolic subtype. Such a profile could be used to predict metabolic treatments that are effective against specific GBM tumor types.

From a therapeutic targeting perspective, numerous metabolic inhibitors have been used in many disease contexts and could be tailored to specific GBM metabolic groups. Classical examples include gemcitabine and fluorouracil, which suppress nucleotide metabolism and nucleic acid synthesis (2). More recent examples that we and others are investigating include the FDA-approved inosine monophosphate dehydrogenase inhibitor known as mycophenolate mofetil (MMF (31)), which is used to block purine synthesis in autoimmunity and is under investigation in GBM (NCT04477200). This may be an especially effective novel therapy in the Base/Carb and Nuc/Ox subgroups, which are also enriched for nucleobases and nucleotides. GBMs in the AA subtype might be treated with amino acid-targeted approaches such as the glutamine antagonist JHU-083, asparaginase, or a methionine-restricted diet (32-34). Lipid metabolism can be targeted by etomoxir or statins and might be an effective approach against GBMs rich in lipids, which likely also encompass Base/Carb and Nuc/Ox GBMs (35).

Our data further suggest that metabolic phenotypes in patients could mediate resistance to standard therapies. Radiotherapy, a standard treatment for GBM, causes DNA damage and oxidative stress. This is notable as we observed an enrichment of metabolites associated with ascorbate metabolism in recurrent GBM compared to primary GBM. Notably, oxidative stress can be targeted by pharmacological ascorbate, which is has shown early promise in patients (3). It could be speculated that ascorbate treatment may be most effective in Nuc/Ox GBMs and recurrent GBM, which is likely to have higher levels of oxidative stress.

While our analyses define subsets of human GBM by metabolite *levels*, metabolic pathway *activity* remains to be defined across subtypes. This can be accomplished by measuring the accumulation of an isotope tracer (for example, ^13^C-glucose) into downstream intermediates. Stable isotope tracing is feasible in both preclinical animal models and cancer patients (36, 37) and could potentially be used in this endeavor. Finally, determining the molecular mechanisms of these phenotypic differences, and how they contribute to tumor progression and therapy resistance, across subtypes *in vitro* and in preclinical animal models will be critical to translation into clinical care. Altogether, these metabolic analyses suggest that gliomas can be grouped into distinct survival groups by metabolite levels and could lay the groundwork to begin developing novel therapeutic strategies for glioma patients.

## Materials and Methods

### Patients, tissue collection and storage

For more than 10 years, the Neurosurgery Department at the University of Michigan has processed and stored resected brain tumor samples not needed for clinical use to facilitate future research endeavors. All samples in this brain tumor bank undergo quality assurance by a clinical neuropathologist to estimate viability and tumor content. Due to the need for banked tissue, patients who only underwent diagnostic biopsy rather than tumor resection are not included in this analysis. Among the types of tissues collected was flash-frozen brain tumor tissue appropriate for metabolomic analysis. Clinical information linked to these samples was abstracted from the medical record under an IRB-approved research protocol.

### Sample preparation

Frozen tissue specimens were homogenized in cold (−80 °C) 80% methanol. Soluble metabolite fractions were separated from insoluble homogenate by centrifugation and dried by speedvac at volumes normalized to equal tissue weights. Dried metabolites were then reconstituted in 1:1 methanol:water for LC-MS.

### Liquid chromatography-mass spectrometry

Metabolite extracts were analyzed using an Agilent Technologies Triple Quad 6470 LC-MS/MS system consisting of the 1290 Infinity II LC Flexible Pump (Quaternary Pump), the 1290 Infinity II Multisampler, the 1290 Infinity II Multicolumn Thermostat with 6 port valve and the 6470 triple quad mass spectrometer. Agilent MassHunter Workstation Software LC/MS Data Acquisition for 6400 Series Triple Quadrupole MS with Version B.08.02 was used for compound optimization, calibration, and data acquisition. Chromatographic separation of compounds is as described (21). Data were pre-processed with Agilent MassHunter Workstation QqQ Quantitative Analysis Software (B0700). For all compounds, the extracted ion chromatograms and mass spectra were manually inspected for sample quality and consistent peak integrations. To validate findings, a second set of samples from patients of an independent cohort (9, 20) was assessed by mass spectrometry by Metabolon.

### Statistical analysis

Descriptive statistics were used to characterize baseline patient and treatment characteristics. Univariable Cox proportional hazard models were used to estimate the association between clinical factors (age and year of diagnosis, performance status, MGMT methylation status, extent of resection, and gender) and overall survival. Kendall’s tau for censored data was used to rank the correlation between metabolites and overall survival (38).

### Metabolomic Analysis

Unsupervised hierarchical clustering, heat map generation, PLS-DA and metabolite set enrichment analysis (MSEA) were performed using MetaboAnalyst 5.0 (27). Processed peak intensities were normalized by the median of all samples, log-transformed and used to generate metabolite-based patient groups by unsupervised hierarchical clustering. Metabolic subtypes were defined as the three largest patient clusters encompassing all GBM samples. Metabolites representing each subtype were identified from the three largest clusters covering all metabolites detected. Network analysis was performed on metabolites representing each subtype and using the joint gene-metabolite interaction network module. Metabolite-gene associations with high confidence were retrieved from STITCH (39). MSEA was performed using metabolites that differed by more than 50% between primary vs. recurrent GBMs with pathways defined by the Kyoto Encyclopedia of Genes and Genomes (https://www.genome.jp/kegg).

### Reduction of LC-MS data with Binner

Metabolite features generated from our metabolomics platform were subjected to Binner quality control analysis (18). Briefly, metabolite features were binned by retention time, and Pearson’s correlation of intensity values were calculated for each feature bin. Isotopes were identified by retention time similarity, correlation, and mass differences. After isotope detection, metabolites in each bin are clustered by correlation coefficients of signal intensities. For each cluster, the highest intensity feature is treated as a neutral mass and iteratively assigned adducts corresponding to the most frequent ions (*e*.*g*., m+H, m+Na). Calculated adducts for each metabolite are searched within the bin based on the m/z of other features in the cluster. Identified adducts were removed from the data set.

### Transcriptional analysis

Htseq quantified RNAseq counts data for 173 TCGA GBM samples were downloaded from the UCSC Xena browser (40). Survival data for these cases were obtained from the TCGA Clinical Data Resource (41). Enrichment scores of each case for specific metabolic genesets were computed with the ssGSEA function in the corto R package (28). We split cases based on positive or negative enrichment scores for the genesets and visualized their survival differences using the Kaplan-Meier method with p-values computed using the log-rank test.

### Other software

All other analyses were performed using GraphPad Prism 8.0.0. Statistical significance between groups was determined by unpaired two-tailed t-tests.

## Data Availability

All data produced in the present study are available upon reasonable request to the authors.

## Conflict of interest statement

None of the authors have relevant conflicts of interest to disclose.

## Funding statement

A.J.S. was supported by the NCI (F32CA260735).

L.O.C. was supported by the NCI (T32CA140044) and the NIAID (T32AI007413).

S.A.C. was supported by the Rogel Cancer Center Core Grant and the Dr. Frank Limpert Career Development Fund at the University of Michigan Rogel Cancer Center.

D.O. was supported by the NCI (R01CA226527).

J.E.I. was supported by the NCI (K99/R00CA218869 and R21CA242221).

P.C. was supported by the NINDS (R01NS110838 and R21NS090087), American Cancer Society grant RSG-11–029-01, and Bankhead-Coley Cancer Research Program.

A.R. was supported by the NCI (R37CA214955), MIDAS PODS grant, MICDE Catalyst grant, and institutional startup funds from the University of Michigan Ann Arbor.

C.A.L. was supported by the NCI (R01CA244931) and UMCCC Core Grant (P30CA046592).

D.R.W. was supported by the NCI (K08CA234416 and R37CA258346), Cancer Center Support Grant P30CA46592, Damon Runyon Cancer Foundation, Ben and Catherine Ivy Foundation and the Sontag Foundation.

---

**Supplemental Figure 1.**
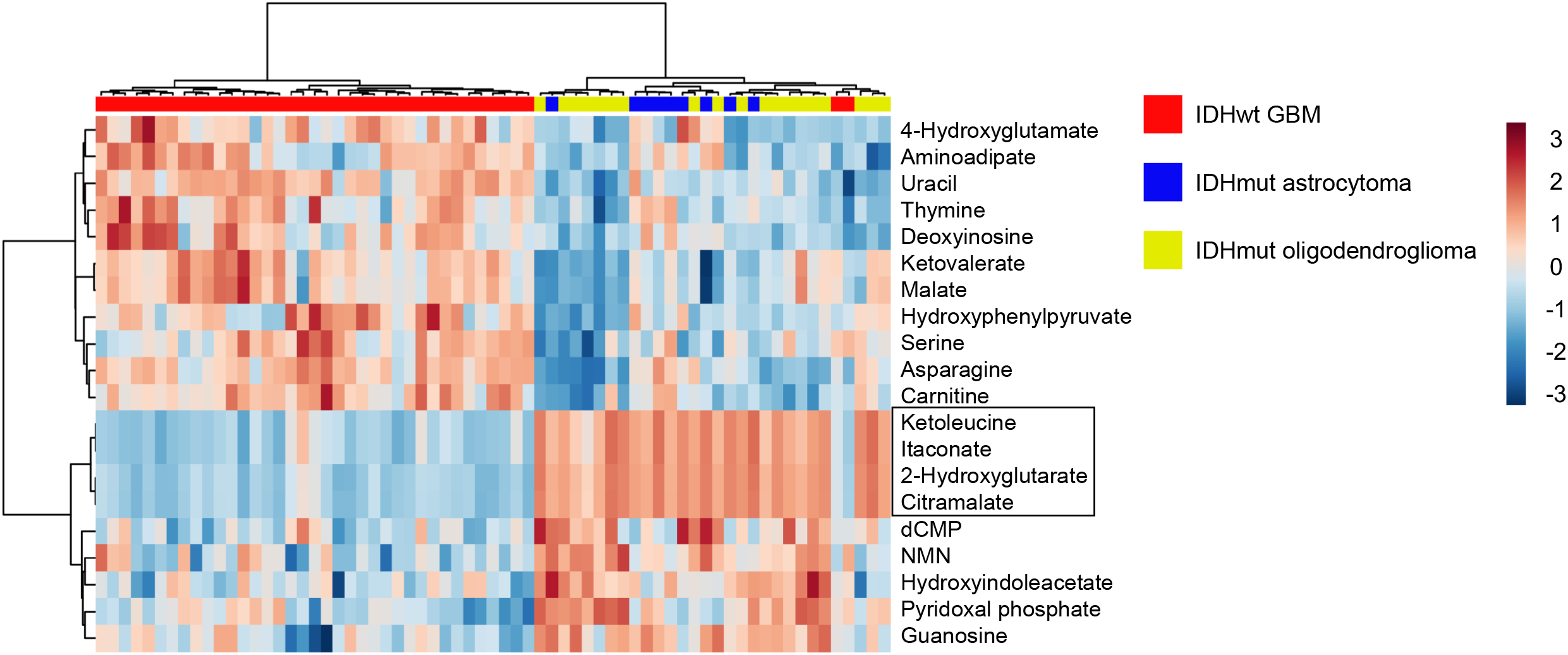
Initial clustering of pre-reduction metabolomics data. Metabolite levels of tumors from glioma patients were measured by LC-MS, and metabolomics data were then subjected to unsupervised hierarchical clustering. Color scale indicates log-transformed values of data points after normalization to the median AUC of each compound. Compound overlaps highlighted in box were identified and corrected using Binner software.

**Supplemental Figure 2.**
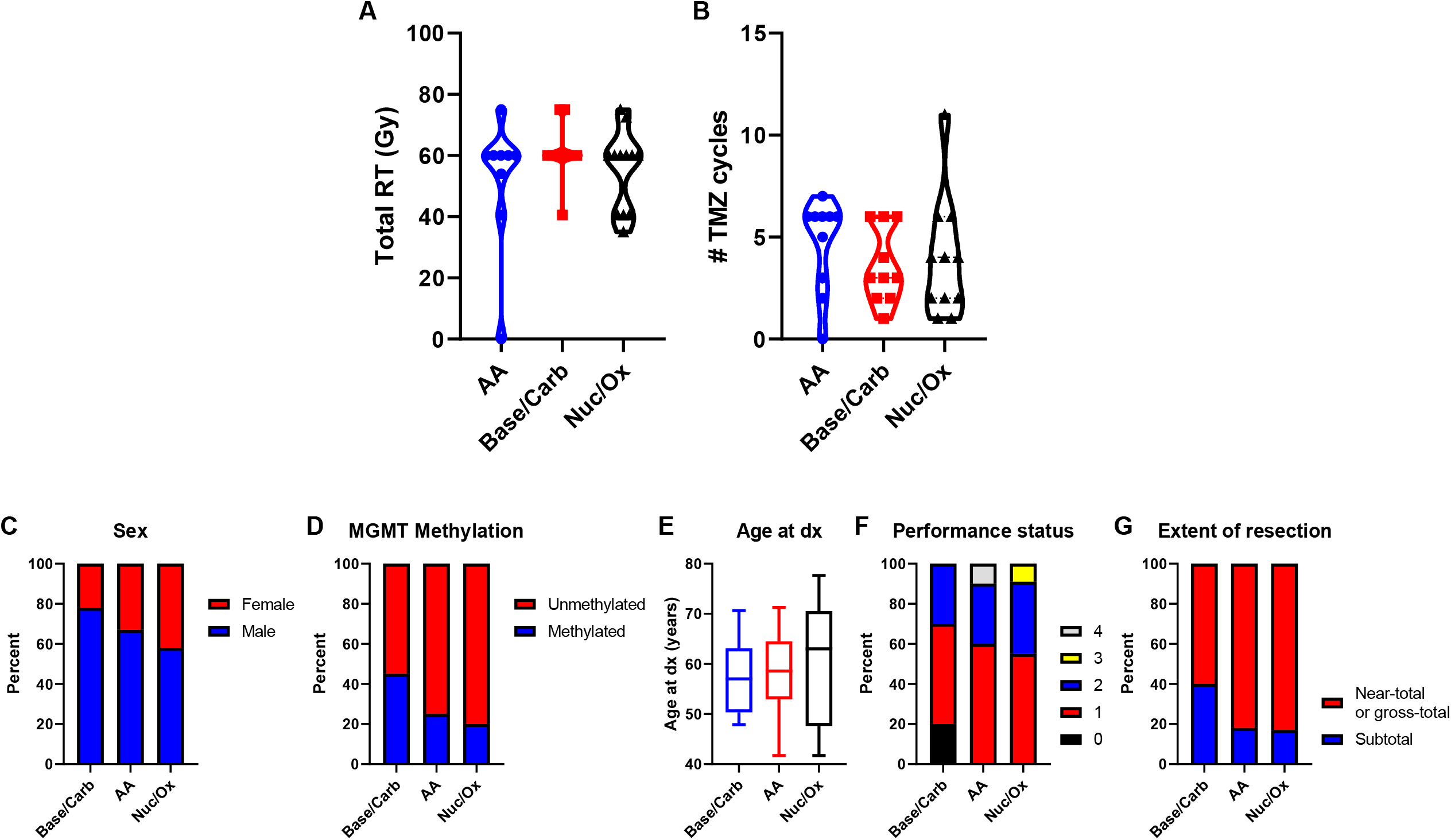
Treatment regimens are similar across metabolic GBM subgroups. *A-B*, Total treatments of RT and TMZ in GBM patients within each metabolic subgroup. *C-G*, Distributions of clinical factors for patients with GBMs in the indicated subtypes.

**Supplemental Figure 3.**
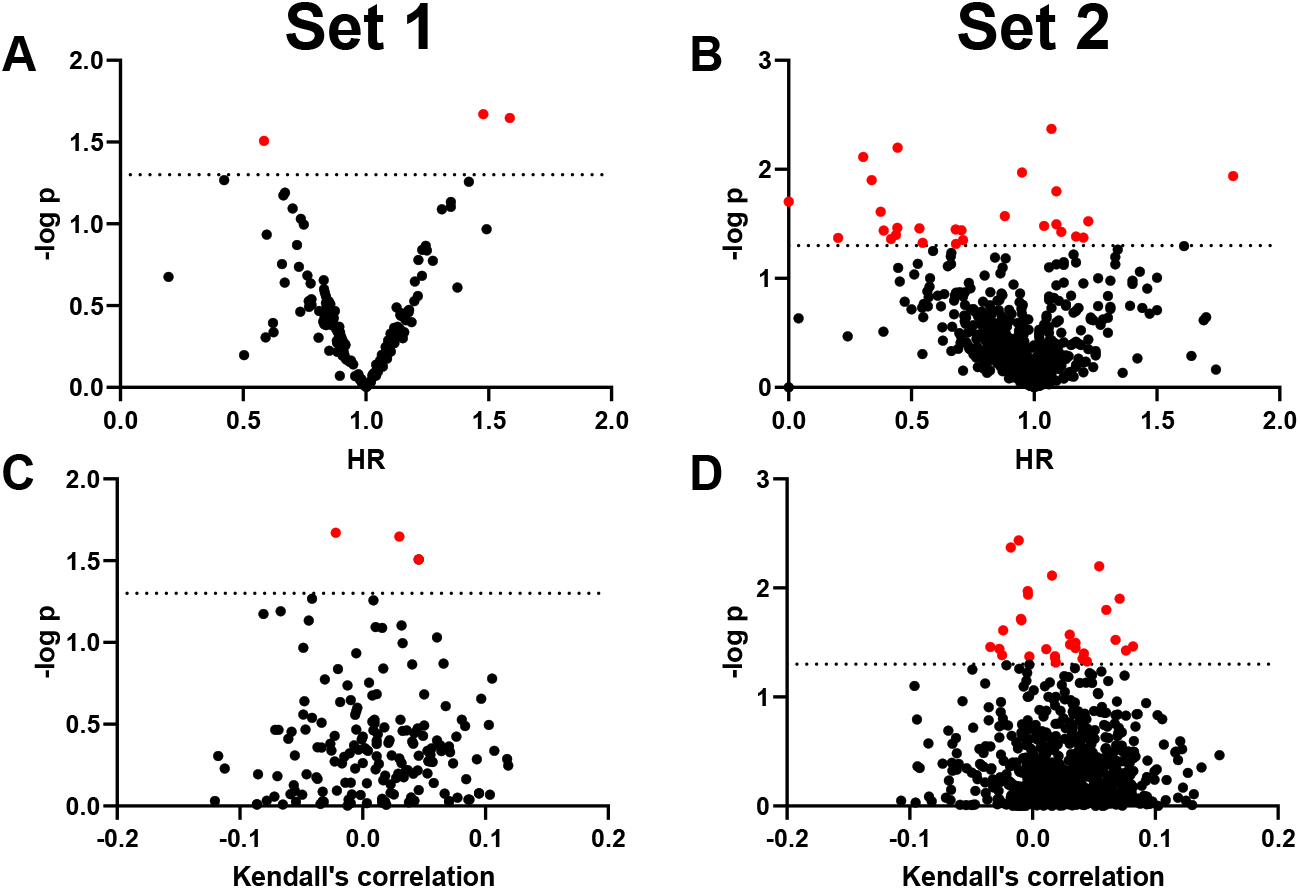
Volcano plots of metabolites in GBMs from set 1 or set 2 (validation cohort) with p-value vs. either hazard ratio (HR) or Kendall’s correlation. Red data points indicate metabolites with p-values below 0.05. No metabolites with p<0.05 in both datasets were identified.

**Supplemental Table 1:** Univariate predictors of survival in GBM patients. Univariable Cox proportional hazard models were used to estimate associations between the indicated clinical factors and overall survival.

|  | HR | Lower | Upper | p | C index |
| --- | --- | --- | --- | --- | --- |
| Age at diagnosis | 1.03 | 1.00 | 1.07 | 0.0465 | 0.6398 |
| Performance status | 1.38 | 0.66 | 2.91 | 0.3890 | 0.5633 |
| MGMT promoter methylation | 0.53 | 0.24 | 1.15 | 0.1093 | 0.5686 |
| Extent of resection | 1.04 | 0.46 | 2.34 | 0.9174 | 0.5000 |
| Sex | 1.36 | 0.64 | 2.89 | 0.4179 | 0.5288 |

## Notes

### Competing Interest Statement

The authors have declared no competing interest.

### Author Declarations

Institutional Review Board of University of Michigan Medical School (IRBMED) gave ethical approval for this work.

